# Enhanced Discrimination of *Clostridioides difficile* Transmission Using Whole Genome Sequencing and *In-Silico* Multi-locus Variable Number Tandem Repeat Analysis

**DOI:** 10.1101/2024.07.22.24310821

**Authors:** Alexander J. Sundermann, Emma G. Mills, Vatsala Rangachar Srinivasa, Marissa P. Griffith, Deena Ereifej, Kady D. Waggle, Graham M. Snyder, Daria Van Tyne, Lora Lee Pless, Lee H. Harrison

## Abstract

Whole genome sequencing (WGS) is used to establish genetic relatedness of bacteria and track outbreaks in healthcare settings. While WGS provides sufficient discriminatory power to make inferences about genetic relatedness and transmission for most bacterial species, WGS for *Clostridioides difficile* often fails to provide discriminatory power, even at low single nucleotide polymorphism (SNP) differences. Multi-locus variable number tandem repeat analysis (MLVA), which analyzes rapidly mutating tandem repeat loci has previously been shown to be useful for this purpose for *C. difficile*. We investigated whether *in silico* MLVA can further elucidate genetic relatedness of *C. difficile* clusters identified by short-read WGS but lacking epidemiological links. Potential healthcare-associated toxin-positive *C. difficile* isolates were collected at our hospital from November 2016 to November 2019. Short-read WGS was performed on the Illumina platform to cluster isolates with ≤2 SNPs, and Nanopore long-read sequencing was used to resolve MLVA loci within these clustered isolates. Among 666 isolates, 62 unique patient isolates met the ≤2 SNP criterion and underwent MinION sequencing. Of the 105 pairs with 0-2 SNP differences, 79.0% had a summed tandem-repeat difference (STRD) of 0-5, 10.5% had an STRD of 6-10, and 10.5% had an STRD of 11-20. A significant correlation was found between a lower STRD value and the presence of a unit/procedure epidemiological link within low SNP clusters (OR: 0.45; 95% CI: 0.29, 0.70). Our findings demonstrate that MLVA provides additional genomic discrimination for closely related *C. difficile* isolates identified by WGS, enhancing outbreak investigation precision.

## INTRODUCTION

Whole genome sequencing (WGS) has become the primary approach for establishing genetic relatedness of bacteria. This method enables investigators to detect and track outbreaks of pathogens throughout healthcare settings and the community. Our hospital uses WGS surveillance in combination with machine learning of the electronic health record to identify otherwise undetected outbreaks and determine the most likely transmission route, respectively.^1^ Although we do not use a strict threshold of single nucleotide polymorphism differences (SNPs) to infer transmission, we have generally found that ≤10-15 SNPs is a valid initial threshold for determining genetic relatedness with supportive epidemiological evidence.^1–4^ However, the *Clostridioides difficile* genome is relatively slow to mutate, which results in lower thresholds of SNP distance to establish genetic relatedness.^5^ Accordingly, it is common to not find an epidemiologic link for isolates that appear to be related genetically, even at low SNP thresholds (e.g., ≤2 SNPs), indicating that WGS provides insufficient discriminatory power for this organism.^5,6^

Tandem repeat loci have been referred to as “mutational hotspots” because they are among the most rapidly changing regions of the bacterial genome.^7^ These loci have a propensity for strand-slippage replication as well as recombination mechanisms that lead to a decrease or increase in the number of repeats. Tandem repeats allow bacteria to rapidly adapt to their environment by regulating gene expression through phase variation and are also a useful tool for epidemiologic tracking. Multi-locus variable number tandem repeat analysis (MLVA) was first developed for *Bacillus anthracis*, an organism for which epidemiologically unrelated isolates could not be distinguished based upon molecular epidemiologic methods that were available at the time.^8^ Traditional MLVA was performed by PCR amplification of tandem repeat loci and inferring the number of repeats at each locus based on the fragment size of the PCR product. The summed tandem repeat difference (STRD) between two isolates is calculated by adding the number of repeat differences at each of the MLVA loci, and can be used to make inferences about transmission.^9^ MLVA was replaced by WGS as the cost and availability of WGS have made the technology more accessible. However, using WGS, MLVA theoretically could be performed *in silico* from WGS data.^10^

An MLVA assay for *C. difficile* was previously developed and used by our group and others to assess the relatedness and hospital transmission of *C. difficile* isolates before WGS became widely available.^9,11^ The assay was performed using PCR-based fragment size-based analysis. However, with the wide-spread availability of WGS, an *in silico* approach can be used. A prior study determined that WGS and MLVA were generally concordant for determining *C. difficile* genetic relatedness.^10^ However, the authors found discordance between the two methods, with some isolates having ≤2 SNPs (suggesting transmission) but an STRD ≥10 (suggesting that transmission was unlikely). These results suggested that MLVA may increase discriminatory power among isolates that appear to be highly related by WGS.

In using routine WGS surveillance for identifying otherwise unrecognized outbreaks of *C. difficile* in our hospital, we have identified genetically related clusters (≤2 SNPs) for which an epidemiologic link could not be identified.^1^ We therefore investigated whether 1) *in silico* MLVA could provide discriminatory power in addition to WGS and 2) whether this approach could be useful epidemiologically in the setting of genomic surveillance for *C. difficile* transmission.

## METHODS

### Study setting

This study was performed at UPMC Presbyterian Hospital, an adult tertiary care hospital with 699 total beds, 134 critical care beds, and over 400 annual solid organ transplants. Ethics approval was obtained from the University of Pittsburgh Institutional Review Board.

### Isolate collection

Culture-independent diagnostic test-positive *Clostridioides difficile* stool specimens were collected between November 2016 and November 2019, as previously described.^1^ We then performed culture of these specimens for *C. difficile*. Inclusion criteria were hospital admission or observation ≥3 days before the collect date and/or a recent inpatient or outpatient encounter in the 30-days before the collect date.

### Whole genome sequencing

Short-read sequencing was performed on the Illumina NextSeq 500 platform (Illumina, San Diego, CA). Reads were assembled with SPAdes v3.13^12^ annotated with Prokka v1.14,^13^ and multi-locus sequence types (STs) were assigned using PubMLST typing schemes (https://github.com/tseemann/mlst).^14^ Pairwise core genome single nucleotide polymorphisms (cgSNP) differences were calculated using snippy v4.3.0 (https://github.com/tseemann/snippy) within species STs having >2 isolates. SNP distances were also calculated using a split kmer approach with SKA v1.0. Genetically related clusters were assigned using initial SNP cutoffs using hierarchical clustering with average linkage using the minimum SNP value from SKA or Snippy for each pair. Clusters were defined as isolates from >1 patient having ≤2 pairwise cgSNPs. Illumina sequence data are available at NCBI BioProject PRJNA475751.^15^ Long-read sequencing was performed on a MinION device (Oxford Nanopore Technologies, Oxford, United Kingdom). Long-read sequencing libraries were prepared and multiplexed using a rapid multiplex barcoding kit (catalog SQK-RBK004) and were sequenced on R9.4.1 flow cells. Base-calling on raw reads was performed using Albacore v2.3.3 or Guppy v2.3.1 (Oxford Nanopore Technologies, Oxford, UK).

### In silico multi-locus variable number tandem repeat analysis

MinION long-read sequencing approach was used to fully and accurately resolve all MLVA loci.^9^ MinION sequencing generates longer reads, which significantly improves the assembly quality and reduces the number of contigs, thereby enhancing the resolution of MLVA loci. In general, short-read sequencing performs more accurate base-calling compared to long read sequencing methods. Therefore, by combining the accuracy of short reads and coverage of long reads, hybrid assembly methods enabled us to resolve all the MLVA loci across the clustered isolates. Hybrid assemblies were constructed and polished using Flye and Polypolish v.0.5.0, respectively.^16,17^ STRDs were calculated in a pairwise fashion for all isolates in a cluster. Pairwise comparisons were categorized based on their STRD as follows: indistinguishable (STRD = 0), highly related (STRD = 1-2), closely related (STRD = 3-4), possibly related (STRD = 5-10), and unrelated (STRD > 10).^9^

### Transmission analysis

Epidemiological links were determined using methods as previously described.^1^ Briefly, clusters were reviewed for unit commonalities, shared procedures, and common healthcare workers prior to their positive test dates. Additionally, a machine learning algorithm utilizing electronic health record charge codes and healthcare worker data was applied as an adjunct to find other plausible routes of transmission.^1^ These routes were reviewed by infection prevention experts to: 1) identify the most likely transmission route among the possibilities suggested by the ML algorithm, and 2) potentially find additional routes of transmission otherwise missed. Transmission routes were stratified into unit epidemiological commonalities alone and any (unit, procedural, or healthcare worker) epidemiological commonality.

### Statistical Analyses

Regression analysis was performed for the correlation of STRD (as a continuous variable) to determine the presence of any epidemiological link or a unit epidemiological link within *C. difficile* clusters (binary outcomes). The outcome model’s performance was assessed using the Area Under the Curve (AUC) of the Receiver Operating Characteristic (ROC) curve. Additionally, an ANOVA with Tukey’s HSD test was conducted to measure the association between STRD and pairwise SNP distance to determine if there were significant differences in STRD across the different levels of pairwise SNP distance. All analysis was performed using SAS v. 9.4.

## RESULTS

Short-read WGS on the Illumina platform was performed on 666 isolates during the 36- month study period (Figure 1). We first attempted to use Illumina short-read sequencing alone to characterize MLVA loci. However, initial data showed inaccurate calling compared to PCR or long-read methods with only 43.7% of 666 isolates with all 7 loci accurately resolved. To explore the utility of long-read sequencing for MLVA, we sequenced 62 unique patient isolates within 19 clusters (range: 2-10 patients per cluster) that were ≤2 SNPs from one another on the Oxford Nanopore MinION platform. Among the 105 isolate pairs with 0-2 SNP differences, 83 pairs (79.0%) had an STRD of 0-5, 11 pairs (10.5%) had an STRD of 6-10, and 11 pairs (10.5%) had an STRD of 11-20.

**Figure 1.**
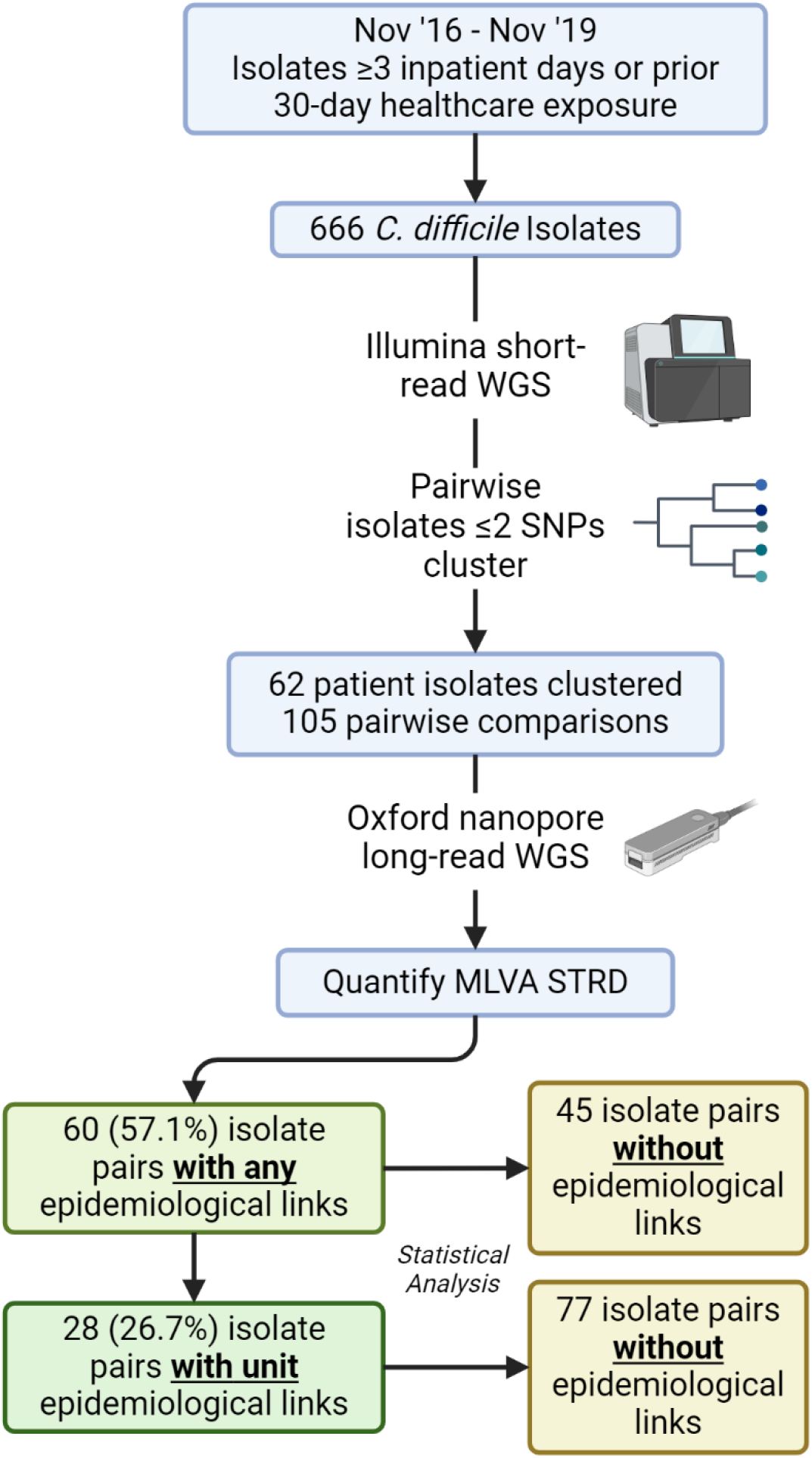
Process and pipeline for multilocus variable number tandem repeat analysis (MLVA) summed tandem repeat difference (STRD) analysis

Overall, 60 (57.1%) isolate pairs had any identified epidemiological link, while 28 (26.7%) had a unit-only epidemiological link. Logistic regression analysis found no significant association between MLVA STRD and the presence of any epidemiological link within these clusters (OR: 0.94; 95% CI: 0.85, 1.04). However, regression analysis revealed a significant correlation between a lower MLVA value and the presence of a unit-based epidemiological link within low SNP clusters (OR: 0.45; 95% CI: 0.29, 0.70) (Figure 2) i.e., for each incremental increase of 1 in STRD, the odds of having a unit-based epidemiological link decrease by 55%. The AUC for the ROC curve was 0.80, indicating good discriminative ability of the model. The ANOVA analysis revealed significant differences in STRD when comparing pairwise SNP distances. Specifically, there was a significant difference in STRD between 0 and 1 SNP pairwise comparisons (p < 0.05; 95% CI: 2.6, 6.2) and between 0 and 2 SNP pairwise comparisons (p < 0.05; 95% CI: 3.5, 7.5). However, there was no significant difference in STRD between 1 and 2 SNP pairwise comparisons (p > 0.05; 95% CI: -1.4, 3.6]).

**Figure 2.**
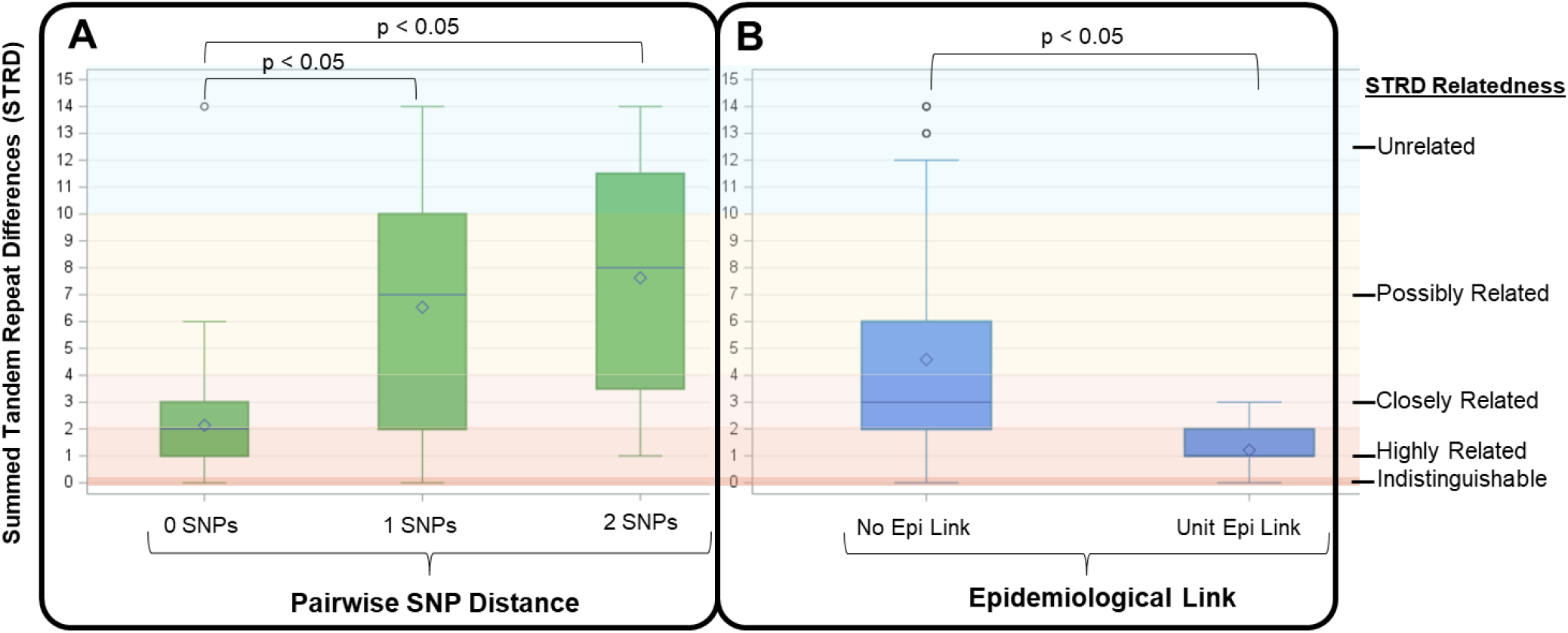
<u>Panel A</u>: Summed tandem repeat difference (STRD) for each pairwise SNP distance (0, n=72 pairs; 1, n=17 pairs; 2, n=16 pairs); <u>Panel B</u>: Relationship between STRD for presence of a unit/procedure epidemiological link (n=28 pairs) versus no epidemiologic link (n=75 pairs), among within ≤2 SNP *C. difficile* clusters.

## DISCUSSION

In this study, we demonstrate that MLVA provides additional genomic discrimination in *C. difficile* isolates that are closely related via WGS. Using this method, many *C. difficile* isolates in SNP-defined clusters without any epidemiological links were removed from clusters when an additional lower STRD threshold was applied. This application may enable hospital epidemiologists to better prioritize outbreaks investigations that have lower STRDs, thus directing investigation resources more effectively towards suspected outbreaks where transmission may truly be occurring. Our data suggest that an STRD of approximately 3 would be a reasonable cutoff, although prospective validation of this approach is required.

Previously, the generation of *in silico* MLVA data was not feasible for routine practice due to the need for multiple pipelines and the associated duplicative costs. Our analysis shows that MLVA cannot be accurately determined via short-read sequencing alone; instead, a long-read sequencing approach is required. Institutions currently performing WGS surveillance should consider incorporating this method into their *C. difficile* investigations. Going forward, our WGS surveillance program has adopted the approach of performing long-read WGS on short-read WGS clusters to better elucidate possible transmission for isolate pairs with small STRDs.

There are limitations to our study. First, we did not sample asymptomatic carriers who could act as transmission sources. Second, although we conducted an exhaustive investigation into transmission routes, there may have been transmissions that were unidentified through our electronic health record review. Nonetheless, here we demonstrate that MLVA has the ability to separate closely related *C. difficile* isolates defined by WGS without epidemiological links. Implementing *in silico* MLVA into our outbreak detection pipeline can better direct infection prevention resources and guide interventions to halt transmission, thereby improving patient safety.

## Data Availability

Illumina sequence data are available at NCBI BioProject PRJNA475751.

https://www.ncbi.nlm.nih.gov/bioproject/475751

## ACKNOWLEDGMENTS

We thank Jane Marsh for her contributions to the paper.

## REFERENCES

1. Sundermann AJ, Chen J, Kumar P, et al. Whole-Genome Sequencing Surveillance and Machine Learning of the Electronic Health Record for Enhanced Healthcare Outbreak Detection. Clin Infect Dis. 2022;75(3):476–482. doi:10.1093/cid/ciab946

2. Marsh JW, Mustapha MM, Griffith MP, et al. Evolution of Outbreak-Causing Carbapenem-Resistant Klebsiella pneumoniae ST258 at a Tertiary Care Hospital over 8 Years. mBio. 2019;10(5):e01945–19. doi:10.1128/mBio.01945-19

3. Sundermann AJ, Babiker A, Marsh JW, et al. Outbreak of Vancomycin-resistant Enterococcus faecium in Interventional Radiology: Detection Through Whole-genome Sequencing-based Surveillance. Clin Infect Dis. 2020;70(11):2336–2343. doi:10.1093/cid/ciz666

4. Sundermann AJ, Chen J, Miller JK, et al. Outbreak of Pseudomonas aeruginosa Infections from a Contaminated Gastroscope Detected by Whole Genome Sequencing Surveillance. Clin Infect Dis. 2021;73(3):e638–e642. doi:10.1093/cid/ciaa1887

5. Miles-Jay A, Young VB, Pamer EG, et al. A multisite genomic epidemiology study of Clostridioides difficile infections in the USA supports differential roles of healthcare versus community spread for two common strains. Microb Genom. 2021;7(6):000590. doi:10.1099/mgen.0.000590

6. Eyre DW, Cule ML, Wilson DJ, et al. Diverse sources of C. difficile infection identified on whole-genome sequencing. N Engl J Med. 2013;369(13):1195–1205. doi:10.1056/NEJMoa1216064

7. Zhou K, Aertsen A, Michiels CW. The role of variable DNA tandem repeats in bacterial adaptation. FEMS Microbiol Rev. 2014;38(1):119–141. doi:10.1111/1574-6976.12036

8. Keim P, Price LB, Klevytska AM, et al. Multiple-locus variable-number tandem repeat analysis reveals genetic relationships within Bacillus anthracis. J Bacteriol. 2000;182(10):2928–2936. doi:10.1128/JB.182.10.2928-2936.2000

9. Marsh JW, O’Leary MM, Shutt KA, et al. Multilocus variable-number tandem-repeat analysis for investigation of Clostridium difficile transmission in Hospitals. J Clin Microbiol. 2006;44(7):2558–2566. doi:10.1128/JCM.02364-05

10. Eyre DW, Fawley WN, Best EL, et al. Comparison of multilocus variable-number tandem-repeat analysis and whole-genome sequencing for investigation of Clostridium difficile transmission. J Clin Microbiol. 2013;51(12):4141–4149. doi:10.1128/JCM.01095-13

11. van den Berg RJ, Schaap I, Templeton KE, Klaassen CHW, Kuijper EJ. Typing and subtyping of Clostridium difficile isolates by using multiple-locus variable-number tandem-repeat analysis. J Clin Microbiol. 2007;45(3):1024–1028. doi:10.1128/JCM.02023-06

12. Bankevich A, Nurk S, Antipov D, et al. SPAdes: a new genome assembly algorithm and its applications to single-cell sequencing. J Comput Biol. 2012;19(5):455–477. doi:10.1089/cmb.2012.0021

13. Seemann T. Prokka: rapid prokaryotic genome annotation. Bioinformatics. 2014;30(14):2068–2069. doi:10.1093/bioinformatics/btu153

14. Jolley KA, Maiden MCJ. BIGSdb: Scalable analysis of bacterial genome variation at the population level. BMC Bioinformatics. 2010;11:595. doi:10.1186/1471-2105-11-595

15. ID 475751 - BioProject - NCBI. Accessed June 12, 2024. https://www.ncbi.nlm.nih.gov/bioproject/475751

16. Wick RR, Holt KE. Polypolish: Short-read polishing of long-read bacterial genome assemblies. PLOS Computational Biology. 2022;18(1):e1009802. doi:10.1371/journal.pcbi.1009802

17. Kolmogorov M, Yuan J, Lin Y, Pevzner PA. Assembly of long, error-prone reads using repeat graphs. Nat Biotechnol. 2019;37(5):540–546. doi:10.1038/s41587-019-0072-8

